# Throat wash as a source of SARS-CoV-2 RNA to monitor community spread of COVID-19

**DOI:** 10.1101/2020.07.29.20163998

**Authors:** Giselle Ibette Lopez-Lopes, Cintia Ahagon, Margarete Aparecida Benega, Daniela Bernardes Borges da Silva, Valéria Oliveira Silva, Katia Corrêa de Oliveira Santos, Lincoln Spinazola do Prado, Fabiana Pereira dos Santos, Audrey Cilli, Claudia Saraceni, Núria Borges da Cruz, Ana Maria Sardinha Afonso, Maria do Carmo Timenetsky, Luís Fernando de Macedo Brígido, the IAL-COVID working group

## Abstract

**Background:** SARS-CoV-2 RNA detection with real time PCR is currently the central diagnostic tool to determine ongoing active infection. Nasopharyngeal and oral swabs are the main collection tool of biological material used as the source of viral RNA outside a hospital setting. However, limitation in swabs availability, trained health professional with proper PPE and potential risk of aerosols may hinder COVID diagnosis. Self-collection with swabs, saliva and throat wash to obtain oropharyngeal wash has been suggested as having comparable performance of regular swab. We performed throat wash (TW) based surveillance with laboratory heath workers and other employees (LHW) at a laboratory research institute.

**Methods:** Consecutive volunteer testing of LWH and external household and close contacts were included. TW self-collection was performed in 5 mL of sterile saline that was returned to original vial after approximate 5 secs of gargle. RNA extraction and rtPCR were performed as part of routine COVID protocols using Allplex (Seegene, Korea).

**Results:** Four hundred and twenty two volunteers, 387 (93%) LHW and 43 (7%) contacts participated in the survey. One or more positive COVID rtPCR was documented in 63 (14.9% CI95 12%-19%) individuals. No correlation was observed between with direct activities with COVID samples to positivity, with infection observed in comparable rates among different laboratory areas, administrative or supportive activities. Among 63 with detected SARS-CoV-2 RNA, 59 with clinical information, 58% reported symptoms at a median of 4 days prior to collection, most with mild disease. Over a third (38%) of asymptomatic cases developed symptoms 1-3 days after collection. Although overall CT values of TW were higher than that of contemporary swab tests from hospitalized cases, TW from symptomatic cases had comparable CTs.

**Conclusions:** The study suggests that TW may be a valid alternative to the detection of SARS-CoV-2 RNA. The proportion of asymptomatic and pre-symptomatic cases is elevated and reinforces the need of universal precautions and frequent surveys to limit the spread of the disease.

## Introduction

SARS-CoV-2 pandemic continues to expand, occupying its yet mostly unchallenged niche among Human populations. This easily transmitted emergent virus originated as a zoonotic transmission in 2019 (Lu 2020). Although inter-species transmission are occasionally being described (Hossain 2020), human–human transmission is it major route of dissemination (WHO 2020).

Social distancing and related measures are the only currently available tools to block transmission of SARS-Cov-2. Although initially many activities were interrupted, allowing the population to stay home and keep distance from potential sources of infection, some sectors, as public health laboratories, increased their pace in the opposite direction to respond to COVID-19.

The use of real time PCR after reverse transcription of viral RNA (rtPCR) in Brazil and other limited resources countries has been hindered by many factors, and in many settings antibodies tests are being implemented as an alternative to direct viral detection. The timing of antibodies, that in the case of COVID may not follow the expected early IgM/late IgG response pattern (Long 2020), as well as the high proportion of false negative results in the first weeks of infection, especially in rapid tests (Hoffman 2020, Shen 2020, Castro 2020) makes the use of IgM as the sole marker of infectivity a precarious choice. Among with cost, availability of reagents, equipment, specialized personnel and different logistic aspects that constrain RNA testing, one other limitation is sample collection. Validated swabs are scarce in many areas of the country, and attempts to minimize the limitations, as the use of fewer swabs per patient do not solve the problem. Limitations in PPE and safe areas for sample collection add to the current limitations. Self-collection seems an attractive alternative to circumvent this limitation. Self-collection of swab has been shown to be as effective as that collected by a health professional (Wehrhahn 2020), but it still needs the swabs that are limited supplies in many areas. Some studies have investigated the use of either Saliva for SARS-Cov-2 (Azzi 2020, Ceron 2020) or throat wash (TW) to COVID (Saito 2020, Guo 2020). TW had been previously evaluated to SARS-CoV-1 (Wang 2004), with results that suggest a good comparability to the detection using swabs. Direct Saliva and TW thus become interesting alternatives as they do not require swabs to provide material for RNA extraction. We conducted a survey based on TW to access COVID RNA positivity in a large public health laboratory at São Paulo metropolitan area.

## Patients and Methods

During the initial months of the COVID-19 swabs and other collection methods were used by LHW in the institute to identify SARS-Cov-2 RNA in upper respiratory tract, but occasionally throat wash (TW) was alternatively used. Due to limitations in swabs, we organized a TW based surveillance in the institute with laboratory personnel, collaborators and other employees (LHW) and with external contacts to a symptomatic individual from the institute, including households and close friends. Since mid-April self-collection was performed using 5 mL of sterile saline in a 50 mL falcon-like tube, a content that was returned to original vial after approximate 5 secs or more of gargle. Volunteers were oriented to perform the gargle at a secure social distance, preferentially outdoors. 50 mL falcon-like tubes were maintained at approx. 4-8°C before and after collection, and were processed at the next available round of extraction. The extracted RNA (Bio Gene, Quibasa or by automated Extraction, Abbott M2000) was tested for COVID-19 using the commercial kit Allplex (Seegene, Korea), which is based on the Charité protocol (Corman, 2020). Samples with amplification in the three viral targets (E, RdRp and N) were considered positive. As recommended for the Influenza assay, Human RNAse P was used to assess the quality of the sample and the presence of inhibitors. Cycle thresholds (CT) up to 37 were considered valid. Although most reactions include three regions (N, E and RdRp), in some runs, as for confirmation, the CT for the N region was the only available CT. We compared the CT obtained at this survey to results generated from contemporary swab collections, sent as routine testing at the institute, that provide SARS-CoV-2 rtPCR testing to clinical services. At the time of the study, these routine testing was limited only to more severe cases, as those that needed hospitalization. To minimize intra assay variation, we included in the statistical analysis the first documented the N region CT from swabs collected samples evaluated in a same run (PCR plate) or contemporary dates of that a TW analysis. In figure 1 we show the results from all three genomic regions. We used the N region CT values for statistical analysis; Pearson chi square or Fisher exact to compare two categorical variables, Mann Whitney to compare two groups and Kruskal Wallis test for three groups to calculate the two-tailed p-value.

**Figure 1.**
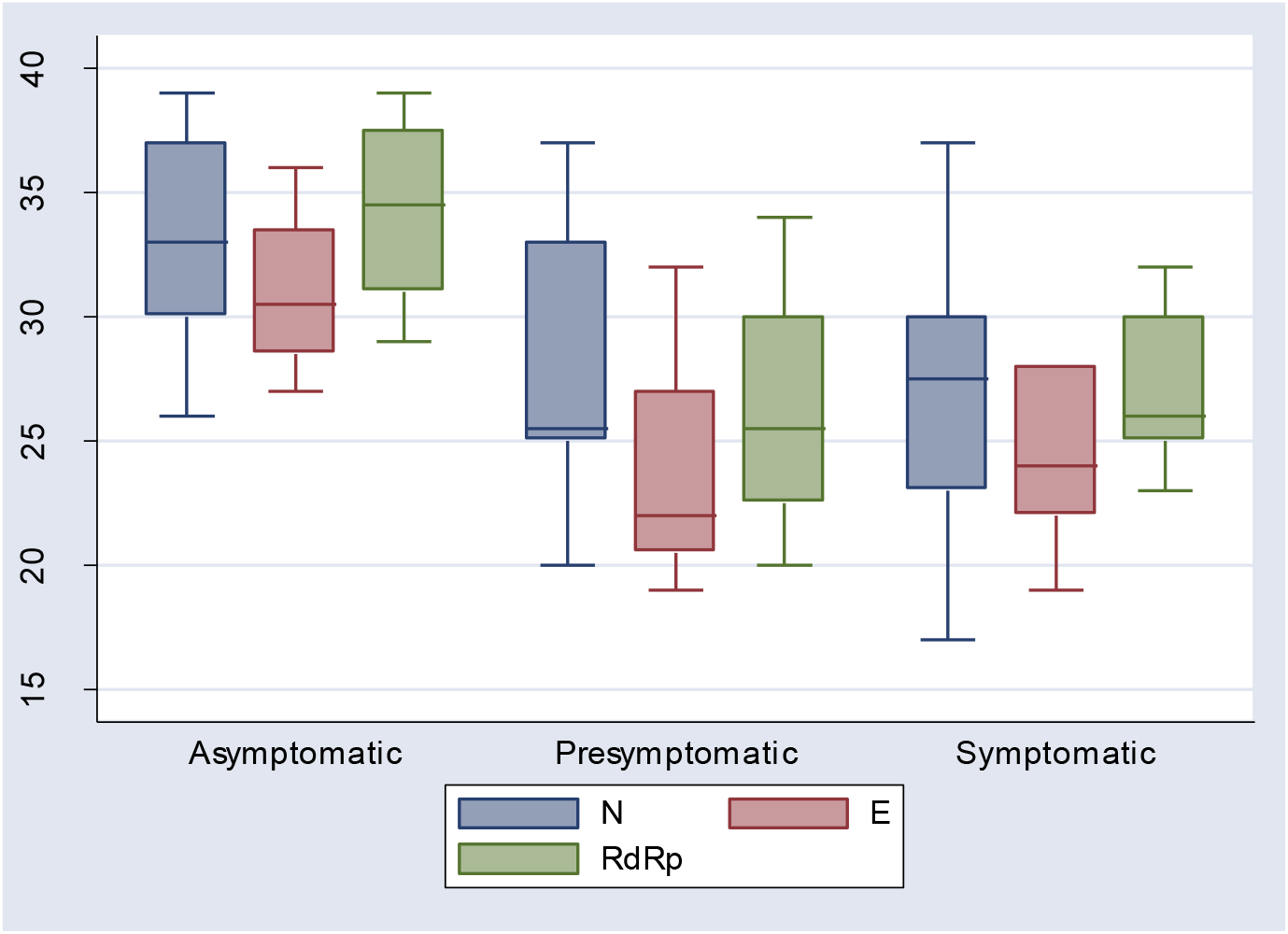
Real time PCR CT values of SARS-CoV-2 according to clinical status depicts a blox plot of rtPCR CT values of SARS-CoV-2 of three viral regions (N, E and Rdrp) of positive, validated tests according to clinical condition at collection For asymptomatic cases, conditions were review after collection to document if symptoms have occurred 2-4 days after collection, these were reclassified as presymptomatic. Only throat wash collections were included. Kluskal Wallis rank test p=0.03.

## Results

From April 15 to July 15, 2020, 387 LHW and 43 contacts (households or close friends) were evaluated for SARS-CoV-2 RNA. Table 1 describes the participants according to rtPCR result. In addition to TW collections, 36 swabs collections (6 positive) were performed at the Institute and 5 at other health services (3/5 positive), as well as 3 self-collected saliva (all negative), registered at the laboratory electronic monitoring system at this period, were also included in this evaluation. Most individuals collected only once (55%), 30% had two collections, 10% three times and 5% collected four or more times. The main reason for repeating a collection was emergence of COVID related symptoms, contact to suspected case after an initial collection in the survey or to evaluate clearance of RNA from samples in positive cases.

**Table 1.** Demographic and work activities characteristics of participants according to SARS-CoV-2 rtPCR result

|  | RNA negative | RNA positive | P |
| --- | --- | --- | --- |
| age | 50 (39-56) | 48 (39-57) | 0.95 |
| Sex ( % females ) | 69.3% | 60.3% | 0.15 |
| Work activity |  |  |  |
| Office | 54 (87.1 %) | 8 (12.9%) |  |
| Biological Lab work | 150 ( 87.2%) | 22 (12.8%) |  |
| other Lab work | 80 (85.1%) | 14 (14.9%) |  |
| outsourced | 43 (87.8%) | 6 (12.2%) | 0.96 |
| COVID related activity |  |  |  |
| Yes | 151 (89.9%) | 17 (10.1) |  |
| no | 298 (87.1%) | 44 (12.9%) | 0.37 |
| All workers | 329 (86.8%) | 50 (13.2%) |  |
| External contactants | 30 (69.8%) | 13 ( 30.2%) | 0.003 |

Four hundred and twenty two health professionals (379, 89.3%) and some external households or close friends contacts (43, 10%) participated in the SARS-CoV-2 rtPCR survey. One or more positive COVID rtPCR test was documented in 63 (14.9% CI95 12%-19%). If only the first collection is considered, the rate is 10.9%. Table 1 shows basic demographic information and the type of work performed, categorized as office, administrative work, contractor from third party firms, and direct laboratory personnel, divided as biological related activities and other laboratory practices that include water and food control, cosmetic and drug safety, chemical analysis, among others. As shown in table 1, there was no association of type of work to positivity rates, which was significantly higher only among external households or friends. When we detail the type of laboratory work according to working with biological COVID samples or not, we also see no difference. If we consider only those working in more risky activities, as opening the biological sample and preparing the extraction of RNA, we also do not have a significant difference (15% vs 12.1%, p=0.63).

Both transmissions within the institute (as during laboratory meetings without strict distancing rules) as well as practically inevitable household events were identified, but we did not evaluate all cases and cannot quantify their importance.

We evaluated in more detail the presence of symptoms at the moment of RNA collection among 59 of those 63 testing positive. A median of 3 symptoms were referred by 58% of the volunteers, at a median of 4 days before the diagnosis. The most common symptoms were fever and anosmia and/or dysgeusia, both reported by 1 in five positive individuals. In 25 (42%) no symptoms were noted at collection, but among 21 of those asymptomatic interviewed again after collection, 8 (38.1%) referred one or more symptom one to three days after collection of sample for RNA diagnosis.

The study did not compare the rate of positivity in paired samples, and only one individual was documented that performed both a swab test (negative) and a positive throat wash collection at a same day. All other swabs included in this study the collection were mostly done at different dates or different heath service. Most (32/36) swabs collections occurred at the first collection.

Considering only the first collection, swabs gave a rate of positivity of 18.8% and TW of 10.4% (p=0.2). Although no direct comparison to a control swab collection was available, the CT observed among positive throat wash collection was compared to that observed in swab analyzed at the time of the study. Using these swabs as a comparator from the same period, throat wash samples tended to have a higher CT (median CT from the N region, 29 for TW and 26 for swabs, p = 0.0002).

Comparing the CT values for the N region of TW collection only, and considering the clinical status at the time of collection, we observed a higher CT for asymptomatic when compared to symptomatic volunteers (33 vs 28, p=0.03). However, if we separate the asymptomatic cases at collection that persisted asymptomatic after collection form those developing symptoms 2-3 days after the RNA test, the latter tended to have a CT value more comparable to that of symptomatic cases (26 vs 28, p= 0.9). There is a significant (p=0.03) difference if the three groups are compared (figure 1).

## Discussion

COVID-19 has been causing a devastating outbreak in countries like Brazil that, albeit having active and free public health system, lacks key elements to respond properly to the COVID challenge, including proper testing capability and consequent contact tracing. Economic and political forces have influenced reopening and other policies, and as end of July 2020 the country experiences the persistence of important morbidity and mortality (WHO 2020). The survey was conducted at a period were the number of cases and death were showing important increase (WHO 2020), possibly reflecting high levels of community transmission, a fact that might had influenced the relatively high rates of infection, 14.9% (CI 95% 11.9%-18.7%) documented in this study. This survey was conducted in an institute that has about 600 professionals and about 100 outsourced workers, including janitors, security and different maintenance teams, along with only part of the students as most were dismissed or did home office. Not only the institute kept working at normal shifts, but in many cases the work burden of some areas even increased to cope with the pandemic demands. Our study included about half of this population, and potential bias in the sampling might had some influence in results. We offered a voluntary collection to all interested, an open consecutive inclusion, but only a few relatives and close friends from some of the symptomatic or infected personnel were included and a proper evaluation of household infection was not possible. The higher proportion of infection in these external individuals possibly reflects inclusion criteria. Symptomatic cases performed tests both within the context of the study as well as directly, especially those that worked in COVID related activities in the institute or in some other health facility. we included all consenting individuals that we identified in this survey both from spontaneous and active search for completeness and to minimize inclusion bias, but some cases were not reached, many asymptomatic did not participate and a few refused participation. The importance of asymptomatic cases has been already observed (Wang 2020) and we can expect that some asymptomatic cases might not have participated in our survey. The findings described here have to take these limitations into account.

Another important point is the selection of throat wash (TW) as the mode of sample collection. The practicality of the method, previous local experience of its use for influenza and mumps, and literature support (Guo 2020, Saito 2020) were the main reasons for this choice. Saliva would also be an option (Azzi 2020), but swabs, the validated method for collection, was simply not available.

Self-collection was performed in 5 mL of sterile saline that was returned to original vial after approximate 5 secs of gargle. The variability of this procedure in each volunteer and the fact that it was done without direct observation might had influence the results. We opt to use a lower volume, 5 mL, whereas the literature suggest up to 20mL (Guo 2020). We felt this volume was adequate and maybe provided a more concentrate wash.

Volunteers from different areas of the institute participated in the study. One or more positive COVID rtPCR was documented in about 15% of case, 11% if only first testing is considered. This rates are higher than the “target levels” of 5 % (WHO 2020) for reopening services. It is also higher than some surveys among Health Care Workers in Italy, 2.3% (Lahner 2020) and US, 5% (Barrett 2020), but lower than that observed for some community outbreaks as in Washington, USA (Kimball 2020), where 30% residents had positive test results. The proportion of asymptomatic cases at this community study (57%) was even higher than that observed in our survey.

No correlation was observed between working with direct activities with COVID samples to a positive result, with infection observed in comparable rates among laboratory, administrative or other supportive activities. Of note, no major difference even if only those involved in more risky activities, as samples preparation before viral inactivation. The fact that these personnel have access to biosafety cabinets and proper PPE may justify these findings, and suggest that transmission is basically from non-bench work related activity. The study so far could not determine the extent of internal versus communal rates of transmission, but internal transmission, as from contact at meeting with someone infected have been documented and are under evaluation.

Close monitoring of fever and other symptoms and swift isolation of cases is a well-known as efficacious and is recommended, but the high proportion of asymptomatic cases demand other measures. Among those with detected SARS-CoV-2 RNA, most symptomatic (51%) had mild disease, with only 4 cases needing hospitalization. Many of those testing positive had no symptoms, with 38% of then developing symptoms 1-3 days after collection. Overall CT values of throat wash were higher than that of contemporary swab tests from hospitalized cases, but TW tests from symptomatic cases showed a comparable CT. The processing of samples to this study, as it was done within the institute, was swift did not had the influence of delays and transport conditions that affected samples collected at the clinical units, but subsequent processing was similar to that of routine samples. If in one side, we tried to show the feasibility and power of TW to identify infections some suggest that they actually may perform better than swabs. As pointed by Ali & Sweeney, samples collected via the oral cavity (such as a throat wash) may yield higher results than naso-pharyngeal swabs test due to some biological characteristics, as high ACE2 receptor expression on the epithelial cells of oral mucosa and the base of the tongue (Xu 2020).

These numbers however expresses a snapshot of a period during the time of a mostly steep slope in cases growth in the region. This is expect to change as pandemic evolves in the region. It however provided an instrument of diminish transmission within the institute, minimizing the impact of labor force shortage and illustrates an alternative in sample collection with potential to be applied in other settings.

## Conclusions

The study suggests that throat wash may be a valid alternative to the detection of SARS-CoV-2 RNA. The proportion of asymptomatic and presymtomatic cases is important and these must not be underestimated in their potential to disrupt essential activities. Asymptomatic cases unrelated to an index symptomatic source are of special concern as current surveillance system may not identify these individuals. Along with the need to reinforce universal precautions against the COVID, regular survey strategies as well as new innovative strategies must be evaluated to guaranteed the continuity of services and minimize the propagation of the virus.

## Data Availability

All anonymized data is available upon request

